# Comparison of the Clinical Implications among Two Different Nutritional Indices in Hospitalized Patients with COVID-19

**DOI:** 10.1101/2020.04.28.20082644

**Authors:** Xuebei Du, Yuwei Liu, Jing Chen, Li Peng, Yalei Jin, Zhenshun Cheng, Harry H.X. Wang, Mingqi Luo, Ling Chen, Yan Zhao

## Abstract

**Background:** Coronavirus disease 2019 (COVID-19) is an emerging infectious disease.It was first reported in Wuhan, China, and then broke out on a large scale around the world.This study aimed to assess the clinical significance of two different nutritional indices in 245 patients with COVID-19.

**Methods:** In this retrospective single-center study, we finally included 245 consecutive patients who confirmed COVID-19 in Wuhan University Zhongnan Hospital from January 1 to February 29. Cases were classified as either discharged or dead. Demographic, clinical and laboratory datas were registered, two different nutritional indices were calculated: (i)the Controlling nutritional status (CONUT) score; (ii) prognostic nutritional index (PNI). We used univariate and multivariate logistic regression analysis to explore the relationship between nutritional indices and hospital death.

**Results:** 212 of them were discharged and 33 of them died. In-hospital mortality was signifcantly higher in the severe group of PNI than in the moderate and normal groups. It was also significantly worse in the severe-CONUT group than in the moderate-, mild-, and normal-CONUT groups. Multivariate logistic regression analysis showed the CONUT score (odds ratio3.371,95%CI (1.124–10.106), p = 0.030) and PNI(odds ratio 0.721,95% CI (0.581–0.896), P=0.003) were independent predictors of all-cause death at an early stage; Multivariate logistic regression analysis also showed that the severe group of PNI was the independent risk predictor of in-hospital death(odds ratio 24.225, 95% CI(2.147–273.327), p=0.010).The CONUT score cutoff value was 5.5 (56.00 and 80.81%; AUC 0.753; 95% CI (0.644–0.862); respectively). The PNI cutoff value was 40.58 (81.80 and 66.20%; AUC 0.778; 95% CI (0.686–0.809); respectively). We use PNI and the COUNT score to assess malnutrition, which can have a prognosis effect of COVID-19patients.

**Conclusion:** The CONUT score and PNI could be a reliable prognostic marker of all-cause deathin patients with COVID-19.

## 1. Introduction

In December 2019, coronavirus disease 2019 (COVID-19), also called severe acute respiratory syndrome coronavirus 2 (SARS CoV-2) occurred in Wuhan, Hubei, China. The disease quickly and violently broke out in other provinces of China and other parts of the world. The numbers of confirmed cases have risen rapidly in the past three months. As of February 20, 2020, it has caused more than 75,000 confirmed COVID-19 cases in China[1]. The epidemiological and clinical characteristics of COVID-19 have been known through previous reports. According to Huang et al., COVID-19 patients had fever, myalgia fatigue and dry cough[2][2][2]. But apart from the typical symptoms, there wereasymptomatic patients and atypical symptoms, such as diarrhea and headache[3].Although most patients were considered to have a good prognosis, elderly patients and patients with chronic underlying diseases or severe clinical symptoms may have a poor prognosis[4]. In addition, high SOFA scores and d-dimers greater than 1 μg / L might serve as risk factors to help clinicians identify patients with poor prognosis early[5].

Nutritional status has attracted more and more attention in various clinical fields. Generally, malnutrition is considered an indicator related to increased morbidity and mortalitys[6]. Therefore, the assessments of early nutritional status of different diseases are important means to identify malnutrition, nutritional risks, and possible benefit to nutritional interventions, as well as a prerequisite for guiding nutritional treatment plans. Ideal nutritional risk screening tools and nutrition assessment methods should accurately reflect the nutritional status of the body and predict the occurrence of malnutrition-related complications. Traditional measurement methods, such as height or body mass index (BMI) or laboratory indicators, such as albumin, prealbumin and total cholesterol levels, have been used to assess the nutritional status of individuals[7]. However, they are limited to reflecting a small portion of nutritional risks and cannot be comprehensive assessement. Interestingly, some studies have shown that malnutrition is linked to the inflammatory response of chronic diseases and the development of autoimmune diseases. At the same time, abnormal nutritional status is related to the prognosis of malignancies and other diseases, such as heart failure, adult spinal deformity and RA[8,9,10,11,12]. Therefore, understanding nutritional status aims to elucidate the prognostic significance of various diseases. This suggests that assessing nutritional status has clinical significance for disease prognosis[6].However, there are currently no standard recognized tools for nutritional risk screening and assessment of nutritional status.Different methods have different effects on the accuracy of predicting the prognosis of the diseases. Therefore, sensitivity, specificity, and ease-to-use tools are often the main basis for clinical nutrition screening and evaluation.Recently, controlling nutritional status (CONUT) score, prognostic nutritional index (PNI) have also been increasingly used as markers of patients’ nutritional status[13,14,15,16].The PNI was calculated by the following formula: 10 × serum albumin (g/dL) + 0.005 × lymphocyte count (per μ L).The lower score means worse nutritional status.The CONUT score is composed of serum albumin, lymphocytes and total cholesterol[11].Higher score means worse nutritional status.

Whether the two screening tools are suitable for assessing the prognosis of patients with COVID-19 remains unknown. Based on the information we have, there is currently no study on the effect of nutritional status on the outcome of COVID-19 patients. Current assessments of the serious consequences and risk factors for death of COVID-19 are not numerous and limitative [19]. Therefore, the purpose of this study was to (i) investigate the relationship between two different nutritional indices and clinical characteristics of patients with COVID-19, and (ii) evaluate the prognostic effect of these indicators as predictors of adverse outcomes in COVID-19 patients

## 2. Methods

### 2.1 Patients

This retrospective cohort study finally included 245 adult patients (≥ 18 years of age) hospitalized at Zhongnan Hospital of Wuhan University. All patients were diagnosed with COVID-19 based on the criteria published by World Health Organization interim guidance.Zhongnan Hospital was the dedicated hospital in Wuhan to treat COVID-19 patients. Hospitalized patients with a diagnosis of COVID-19 in Zhongnan Hospital of Wuhan University were consecutively enrolled in the present study (n=270), from January 1, 2020 to February 20, 2020.A total of 25 patients were excluded: Life support (such as cardiopulmonary resuscitation (CPR), vasopressors, mechanical ventilation, extracorporeal membrane oxygenation (ECMO)) before admission died immediately after admission: N = 2 (8%); lack most baseline data: N = 1 (4%); Transfer to other designated hospitals: N = 10 (40%); Pregnant woman: N = 6 (24%); Under 18 years old: N = 6 (24%). The rest of 245 cases were included in this study.This study which complied with the Declaration of Helsinki (Approval Number 2020054)was approved by the Medical Ethical Committee of Zhongnan Hospital of Wuhan University.

### 2.2 Clinical and Laboratory Data Collection

The demographic data collected age and gender. About baseline characteristics, gastrointestinal symptoms, comorbidities and laboratory test results are from the Electronic Medical Record System of Zhongnan Hospital. Collection of laboratory data includes Heart rate, Respiratory rate, Systolic pressure,Diastolic pressure, Sodium(Na+), Potassium(K+), white blood cell count (WBC), lymphocyte count, hemoglobin, Platelets, serum creatinine, Alanine aminotransferase (ALT), uric acid (UA), albumin, total cholesterol(TC), high density lipoprotein (HDL) cholesterol,Procalcitonin(PCT). Prothrombin time(PT),D-dimer.

### 2.3. Nutritional Indices Selection and Calculation

The study includestwo different nutritional indicators: CONUT score, PNI. The CONUT score, PNI are calculated according to the following formulas: (i) The CONUT score is calculated based on 3 laboratory variables: serum albumin concentration, total cholesterol concentration, and total peripheral lymphocytes cell count;Based on their CONUT scores, patients were divided into four groups: normal-CONUT (0–1), mild-CONUT (2–4), moderate-CONUT (5–8) and severe-CONUT(≥9)[17]; (ii) PNI is 10 x serum albumin value (g / dL) + 0.005 x total peripheral blood lymphocyte count(unit/L);Patients were also divided into threegroups based on their PNI:severe-PNI (PNI < 35,), moderate-PNI (35 ≤ PNI < 38,), and normal-PNI (PNI ≥38) [18].

### 2.4 Statistical Analysis

Analysis was performed using SPSS (version 22.0). Continuous variables were expressed as median (interquartile range, IQR). Spearman rank correlation analysis was used to analyze the correlation between nutritional indicators and selected baseline clinical data. We used the Mann Whitney U test, χ^2^ test, or Fisher’s exact test to compare the differences between categorical groups.Spearman rank correlation analysis was used to analyze the correlation between nutritional indicators and selected baseline clinical datas. Calculated the sensitivity and specificity of the ROC curve to predict hospital mortality and Used Youden to determine the AUC and cutoff value.Univariate and multivariate logistic regression models exploreddeath-related risk factors. Two-way p-values <0.05 are considered statistically significant.

## 3. Results

### 3.1 Clinical Characteristics of Patients

The baseline characteristics of 245 patients in this study was described in Table 1. Of the 245 patients who were included in the study; severe group of PNI (PNI < 35, n = 31), moderate group of PNI (35 ≤ PNI < 38, n = 30), and normal group of PNI (PNI ≥ 38, n = 182). the CONUT score: normal-CONUT (0−1, n = 18), mild-CONUT (2−4, n = 72), moderate-CONUT (5−8, n =58), and severe-CONUT (≥9, n = 7);The severe group of PNI had significantly lower lymphocyte counts, albumin, high density lipoprotein (HDL)and total cholesterol(TC), and signifcantly higher white blood cells(WBC), alanine aminotransferase (ALT), prothrombin time (PT),D-dimer,Procalcitonin(PCT), than the normal and moderate groups. Some observedvariables were different between the severe-CONUT group and the mild-, and normal-CONUT groups (Tables 1, 2).The in-hospital death was signifcantly worse in the severe PNI group (n = 12, 38.71%) than in the moderate-(n = 8, 26.67%) and normal-(n = 13, 7.14%) groups. It was also signifcantly worse in the severe-CONUT group (n = 6, 85.71%) than in the moderate-(n = 12, 20.69%), mild-(n = 6, 8.3%), and normal-CONUT (n = 1, 5.6%) groups (Fig. 1)

**Table 1.** The patient characteristics and Prognostic Nutritional Index values^*^

|  |  |  |  |  |  |
| --- | --- | --- | --- | --- | --- |
| Characteristic | total | normal-PNI | moderate-PNI | severe-PNI | P |
| No.of patients | 243 | 182 | 30 | 31 | - |
| Age | 55 ( 39-67 ) | 51 ( 36-62 ) | 62 ( 54-70 ) | 71(66-78) | <0.001 |
| Male(%) | 113(46.50) | 73(40.11) | 17(56.72) | 23(74.19) | 0.001 |
| BMI | 23.70<br>( 20.94-25.65 ) | 23.43<br>( 20.82-25.30 ) | 24.73<br>(23.74-28.13) | 23.78<br>(20.45-25.36) | 0.003 |
| **Gastrointestinal symptoms** |  |  |  |  |  |
| Anorexia(yes,%) | 55(22.63) | 28(15.38) | 12(40.00) | 15(48.39) | <0.001 |
| Diarrhea(yes,%) | 18(7.40) | 16(8.79) | 1(3.33) | 1(3.23) | 0.183 |
| Vomit(yes,%) | 10(4.15) | 8(4.40) | 1(3.33) | 1(3.23) | 0.718 |
| Nausea(yes,%) | 17(7.00) | 13(7.14) | 3(10.00) | 1(3.23) | 0.607 |
| **Comorbidities** |  |  |  |  |  |
| Cancer(yes,%) | 9(3.70) | 5(2.75) | 3(10.00) | 1(3.23) | 0.242 |
| COPD(yes,%) | 8(3.29) | 5(2.75) | 1(3.33) | 2(6.45) | 0.313 |
| Hypertension<br>disease(yes,%) | 52(21.40) | 27(14.84) | 14(46.67) | 11(35.48) | <0.001 |
| Diabetes(yes,%) | 23(9.47) | 10(5.49) | 10(33.33) | 3(9.68) | <0.001 |
| Coronary heart<br>disease(yes,%) | 18(7.41) | 9(4.95) | 4(13.33) | 5(16.13) | 0.058 |
| **Laboratory findings** |  |  |  |  |  |
| Heart rate<br>beats /min | 84 ( 76-96 ) | 84 ( 76-95 ) | 89(80-103) | 84(70-96) | 0.133 |
| Respiratory rate<br>beats /min | 20 ( 19-21 ) | 20 ( 19-21 ) | 20(20-23) | 20(19-22) | 0.960 |
| Systolic<br>pressure mmHg | 12 ( 115-133 ) | 122 ( 114-132 ) | 125(113-134) | 122(115-140) | 0.673 |
| Diastolic<br>pressure mmHg | 74 ( 67-82 ) | 75 ( 68-82 ) | 73(66-82) | 73(64-82) | 0.654 |
| WBC ,109/l | 4.38<br>( 3.30-6.17 ) | 4.23<br>( 3.11-5.41 ) | 5.36<br>(3.40-9.00) | 7.16<br>(4.71-11.68) | <0.001 |
| Lymphocyte,109/l | 0.94<br>( 0.61-1.20 ) | 1.04<br>( 0.73-1.32 ) | 0.60<br>(0.47-0.80) | 0.44<br>(0.34-0.73) | <0.001 |
| Hemoglobin ,g/dl | 128.0<br>( 117.7-139.3 ) | 130.0<br>( 118.4-139.7 ) | 125.5<br>(119.0-137.3) | 119.9<br>(103.4-144.0) | 0.680 |
| Platelets,109/l | 176<br>( 130-222 ) | 178<br>( 139-221 ) | 175.5<br>(119.3-220.5) | 158<br>(106-242) | 0.538 |
| ALT, u/l | 22 ( 15-35 ) | 20 ( 14-28.25 ) | 31(20-55.3) | 38(20-55) | <0.001 |
| Albumin, g/l | 37.1<br>( 33.9-40.7 ) | 38.8<br>( 36.2-41.8 ) | 33.4<br>(32.2-34.6) | 29.5<br>(27.2-31.3) | <0.001 |
| HDL, mmol/l | 1.00 | 1.05 ( 0.9-1.295 ) | 0.86(0.76-1.15) | 0.71(0.57-1.03) | <0.001 |
|  | (0.81-1.23) |  |  |  |  |
| Uric acid,umol/l | 263.7<br>(210.3-336.7) | 265.8<br>(210.2-344.6) | 251.8<br>(209.9-324.2) | 256.3<br>(171.7-319.8) | 0.696 |
| Creatinine,μmol/l | 67.4<br>(56.3-85.3) | 66.0<br>(54.9-82.9) | 71.5<br>(63.9-87.7) | 71.9<br>(57.7-95.8) | 0.055 |
| Potassium(K+),<br>mmol/l | 3.97<br>(3.58-4.24) | 3.98<br>(3.67-4.26) | 3.86<br>(3.39-4.13) | 3.64<br>(3.21-4.26) | 0.121 |
| Sodium(Na+),<br>mmol/l | 138.05<br>(135.3-140.4) | 138.6<br>(136-140.6) | 135.2<br>(133.4-138.4) | 138.8<br>(134.0-140.4) | 0.110 |
| TC, mmol/l | 3.85<br>(3.31-4.58) | 3.97<br>(3.45-4.70) | 3.69<br>(2.88-4.12) | 3.73<br>(3.02-4.38) | 0.019 |
| PT,s | 12.8<br>(11.9-13.5) | 12.6<br>(11.9-13.2) | 13.0<br>(12.0-14.2) | 13.5<br>(12.8-14.5) | <0.001 |
| D-dimer ng/ml | 208 (127-455) | 166 (109-295) | 317(213-986) | 658(348-3372) | <0.001 |
| PCT,ng/ml | 0 (0.00-0.10) | 0 (0.00-0.00) | 0.09(0.00-0.31) | 0.11(0.00-0.59) | <0.001 |
| All-cause death | 33(13.58%) | 13(7.14%) | 8(26.67%) | 12(38.71%) | <0.001 |

COPD, chronic obstructive pulmonary disease; WBC, white blood cells;
ALT, alanine aminotransferase;TC,Total cholesterol; HDL, high density lipoprotein
PT, prothrombin time;PCT,Procalcitonin; CONUT, Controlling nutritional status;
PNI, Prognostic nutritional index; BMI, Body mass index
\*Two patients did not detect lymphocytes or albumin

**Table 2.**
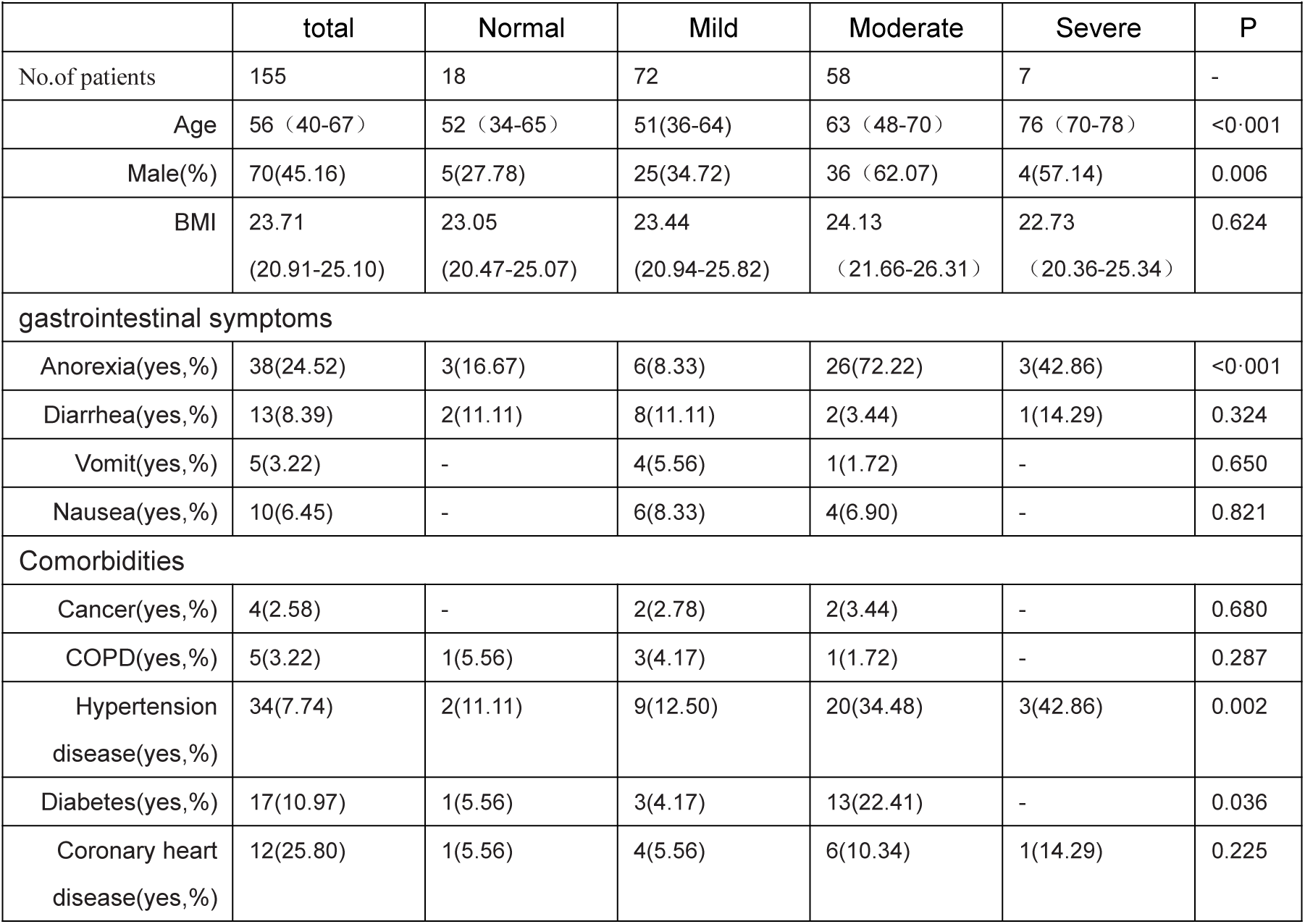
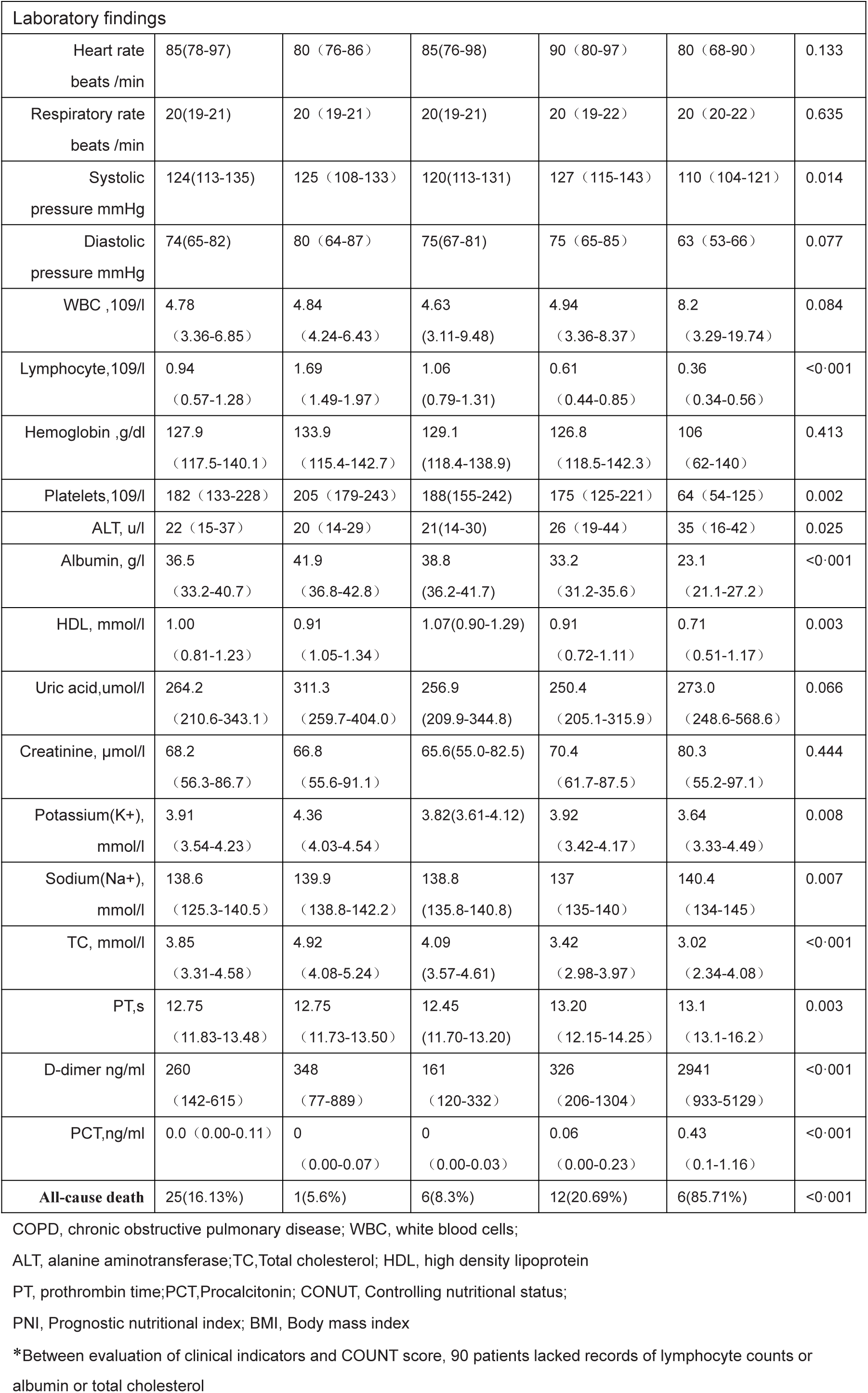
The patients’ characteristics and CONUT score*

**Table 3.** Correlation analysis between variables and nutritional indices

|  |  |  |  |  |
| --- | --- | --- | --- | --- |
| Variables | COUNT |  | PNI |  |
|  | rs | p value | rs | p value |
| Age | 0.363 | <0.001 | -0.582 | <0.001 |
| Male | -0.297 | <0.001 | 0.198 | 0.002 |
| gastrointestinal symptoms |  |  |  |  |
| Anorexia | 0.353 | <0.001 | -0.266 | <0.001 |
| Diarrhea | -0.082 | 0.308 | -0.044 | 0.497 |
| Vomit | -0.015 | 0.851 | -0.043 | 0.502 |
| Nausea | 0.049 | 0.548 | -0.078 | 0.228 |

| Comorbidities |  |  |  |  |
| --- | --- | --- | --- | --- |
| Cancer | 0.020 | 0.803 | -0.103 | 0.108 |
| COPD | -0.068 | 0.401 | -0.051 | 0.426 |
| Hypertension | 0.284 | <0.001 | -0.285 | <0.001 |
| Diabetes | 0.208 | 0.010 | -0.248 | <0.001 |
| Coronary heart disease | 0.149 | 0.063 | -0.126 | 0.050 |
| Laboratory findings |  |  |  |  |
| Heart rate | 0.087 | 0.284 | -0.070 | 0.274 |
| Respiratory rate | 0.083 | 0.307 | -0.158 | 0.014 |
| Systolic pressure | 0.107 | 0.186 | -0.135 | 0.036 |
| Diastolic pressure | -0.144 | 0.074 | 0.050 | 0.439 |
| white blood cell | 0.134 | 0.097 | -0.217 | 0.001 |
| Lymphocyte | -0.719 | <0.001 | 0.654 | <0.001 |
| Hemoglobin | -0.096 | 0.233 | 0.238 | .<0.001 |
| Platelets | -0.277 | <0.001 | 0.093 | 0.148 |
| ALT | 0.235 | 0.003 | 0.281 | <0.001 |
| Albumin | -0.695 | <0.001 | 0.904 | <0.001 |
| HDL | -0.310 | <0.001 | 0.305 | <0.001 |
| TC | -0.557 | .<0.001 | 0.212 | 0.008 |
| Uric acid, | -0.069 | 0.391 | 0.128 | 0.047 |
| Creatinine, | 0.156 | 0.052 | -0.126 | 0.049 |
| K+ | -0.167 | 0.037 | 0.210 | 0.001 |
| Na+ | -0.262 | 0.001 | 0.186 | 0.004 |
| PT | 0.351 | <0.001 | -0.224 | <0.001 |
| D-dimer | 0.419 | <0.001 | -0.548 | <0.001 |
| PCT, | 0.389 | <0.001 | -0.424 | <0.001 |

**Fig. 1.**
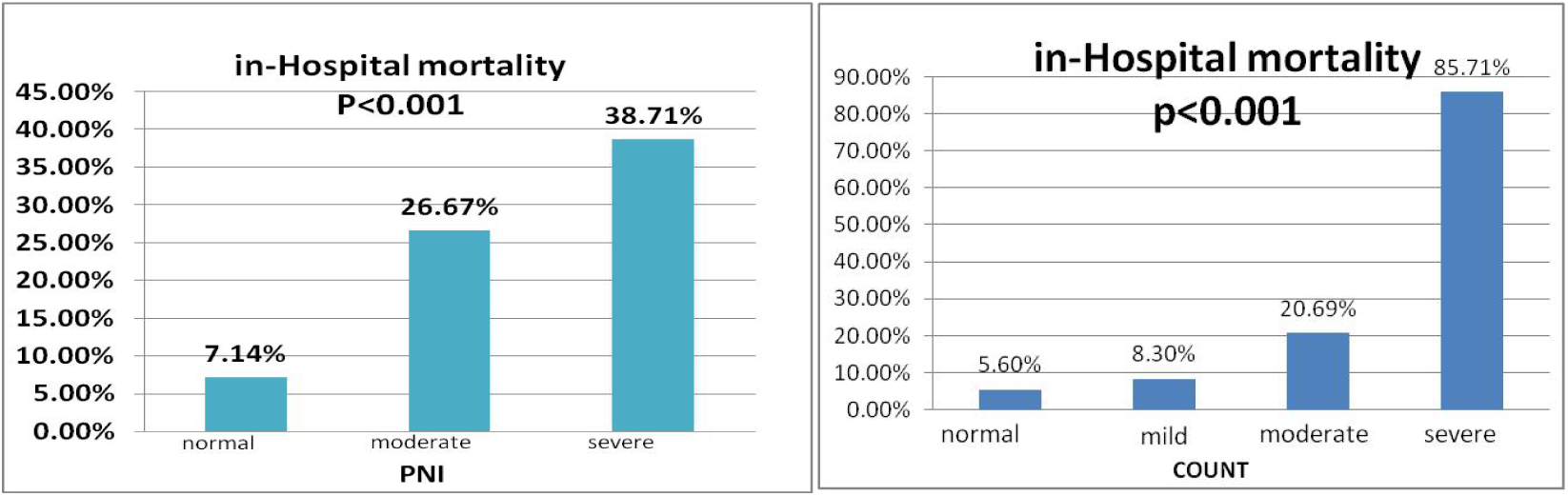
In-hospital mortality of the A PNI, B CONUT score

The median age was 55.0 years, of which 114 patients (46.5%) were male(TabelA). 33 patients died during hospitalization and 212 were discharged(Tabel A). In terms of comorbidities, hypertension was the most common comorbidity(21.2%), followed by diabetes(9.4%) and coronary heart disease(7.4%) (Table A). The most common gastrointestinal symptom before admission was anorexia(22.9%), followed by diarrhea(7.4%) (Table A). In the comparison of gastrointestinal symptoms, we could find that compared with survivors, non-survivors are more prone to anorexia (57.6% vs 17.5%, p <0.001)(Table A).

### 3.2 Correlation between Variables and Nutritional Indices

**Table 2** shows the correlation between nutritional indices and selected characteristics or laboratory variables. The CONUT scores was signifificantly correlated with gender, age, anorexia, hypertension, diabetes, platelets, ALT, HDL, TC, sodium, PT, D-dimer, albumin, lymphocyte count, and PCT. the COUNT score had the highest correlation with lymphocyte, followed by albumin (r = −0.719, and r = = −0.695, all p <0.001). PNI was correlated withage, anorexia, hypertension, diabetes, white blood cell, lymphocyte count, hemoglobin, ALT, albumin, HDL, TC, potassium, PT, D-dimer, and PCT. the PNI had the highest correlation with albumin, followed by lymphocyte count (r = 0.904, and r = = 0.654, all p <0.001).

### 3.3 Factors associated within-hospital death

This study used logistic regression analysis to clarify the factors related to death. In univariate analysis, age, gender, BMI, anorexia, hypertension, diabetes, coronary heart disease, respiratory rate, white blood cell count, ALT, creatinine, PCT,PNI and COUNT were significantly associated with death (Table 4). In the multivariate logistic regression model, we found that the CONUT score (odds ratio3.371,95% CI (1.124–10.106), p = 0.030) and PNI(odds ratio 0.721,95% CI (0.581–0.896), P=0.003) were the independent predictors of all-cause death after adjusting for age, sex, BMI, history of hypertension, history of chronic obstructive pulmonary disease, history of coronary heart disease, respiratory rate, alanine transaminase, creatinine, D-dimer, white blood cell count, Lymphocyte, PCT.A multivariate logistic regression model showed that severe group (classifedin PNI groups) was an independent predictor of all-cause death (odds ratio 24.225,95% CI(2.147–273.327), p=0.010), and in categorical groups of CONUT scorewerenot the independent predictors of all-cause death(Table 5). The CONUT score, which demonstrated sensitivity and specificity for predicting in-hospital death were 56.00% and 80.81% (AUC 0.753; 95% CI (0.644 – 0.862), p<0.001).The cutoff value of the COUNT scorewas 5.5 overall(Fig. 2A).The PNIcutoff valuewas 40.58 overall (81.80and 66.20%, respectively; AUC 0.778; 95%CI (0.686–0.809), p<0.001)(Fig. 2B).

**Table 4.**
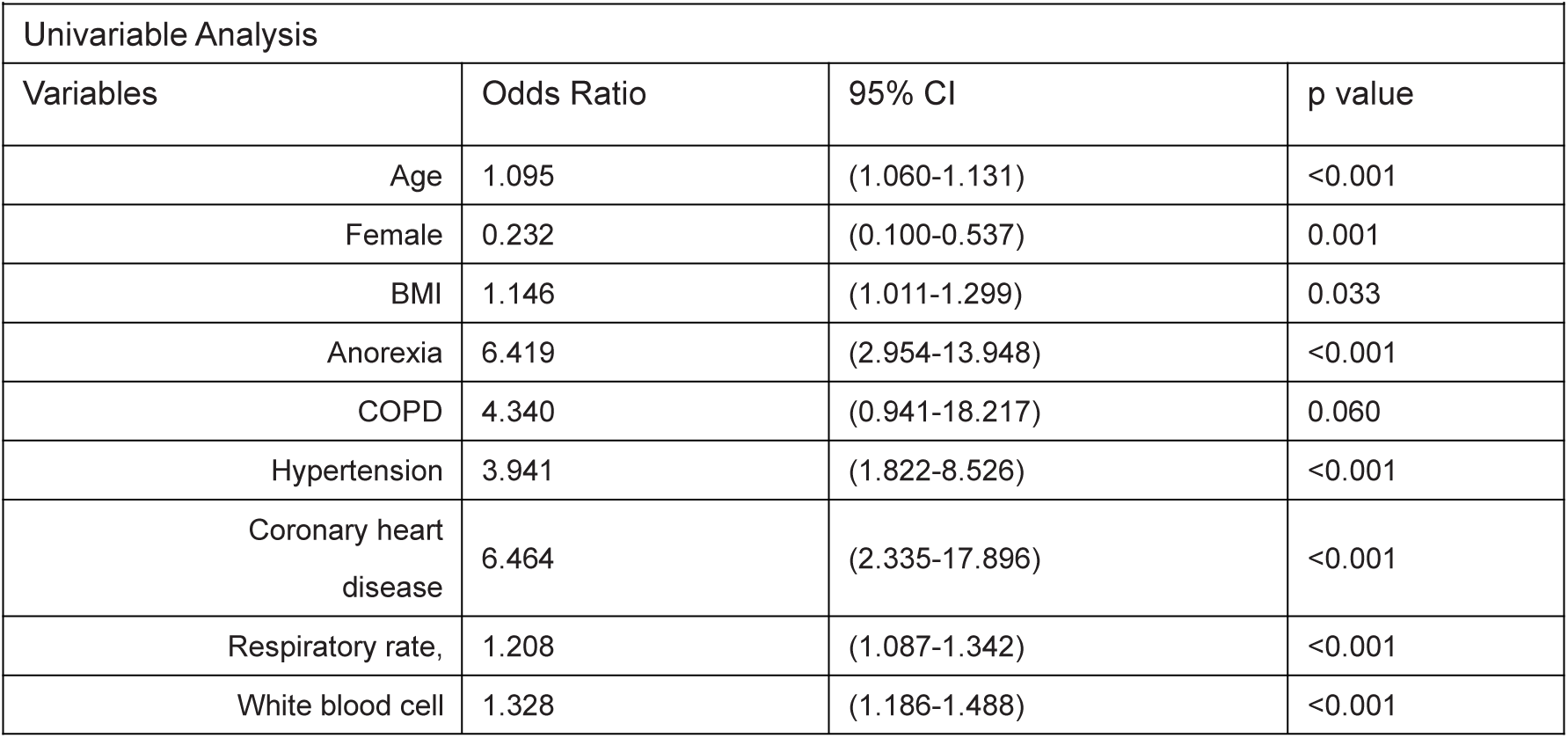
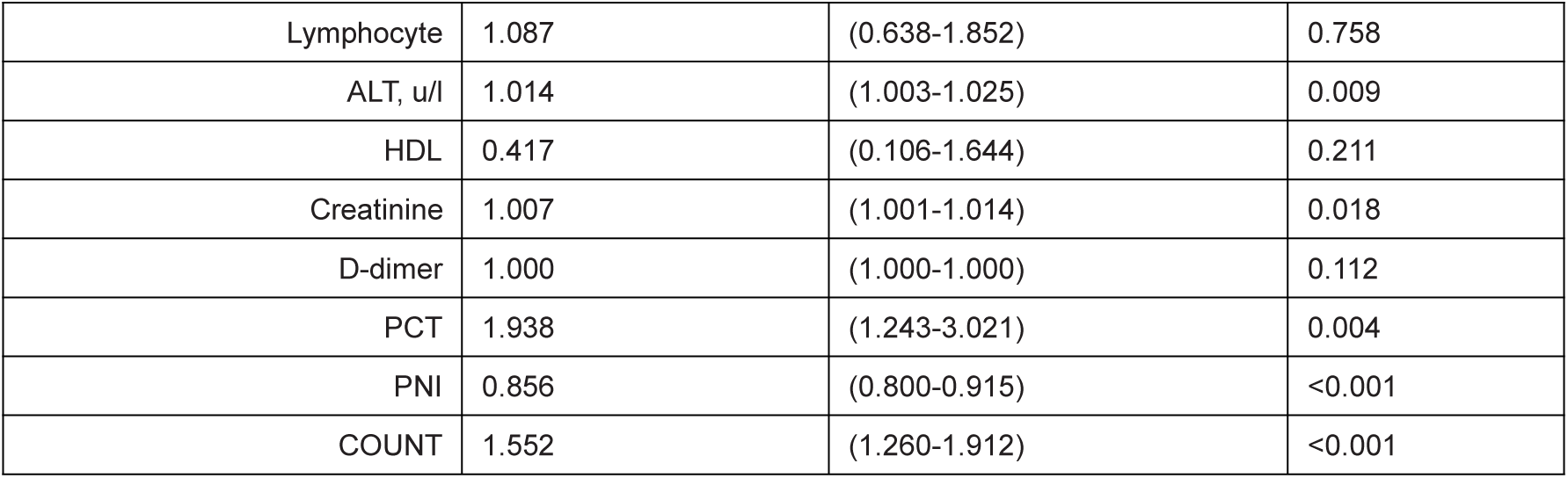
The unadjusted association between baseline variables and all-cause death duringhospitalization

**Table 5.**
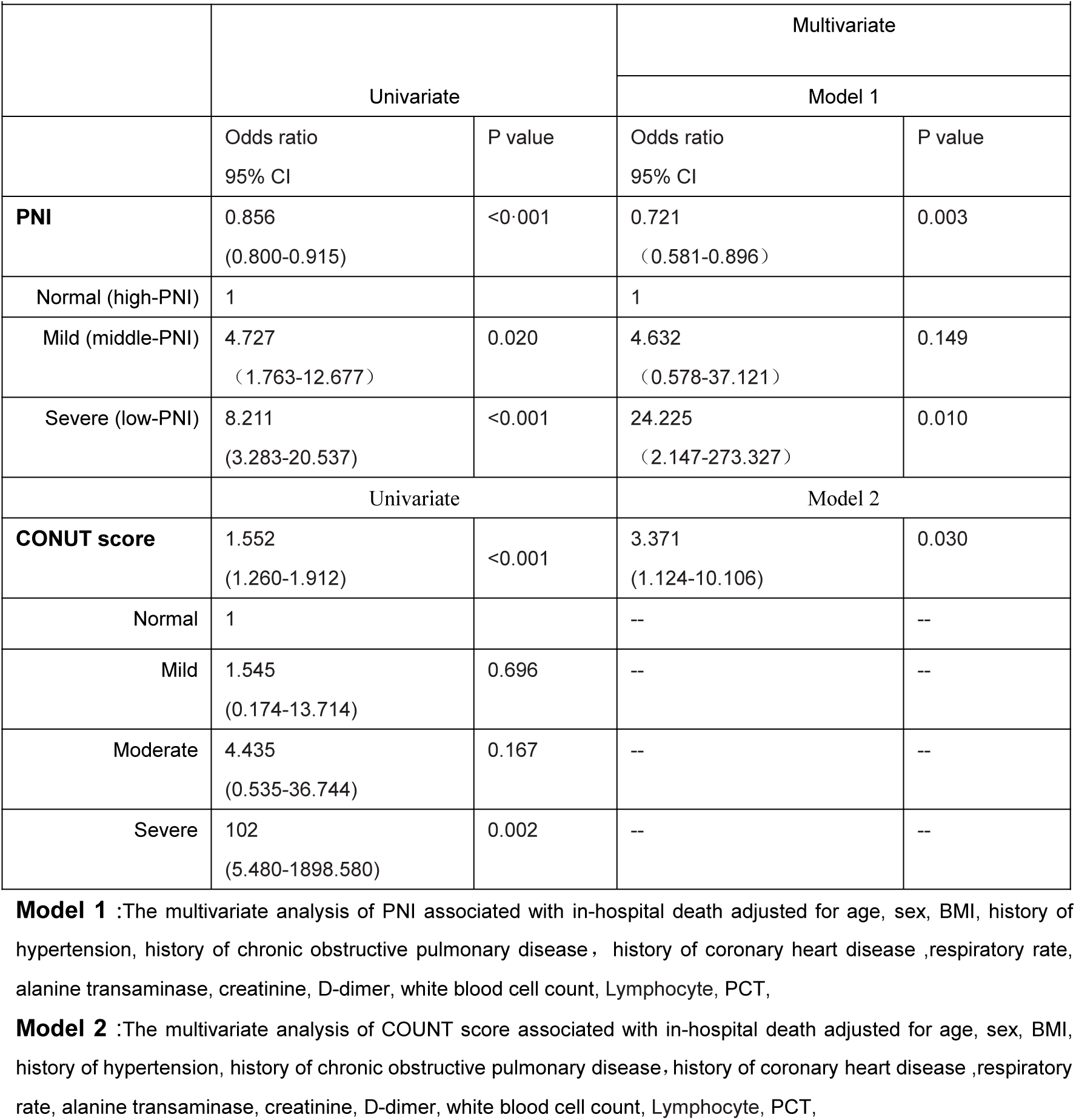
The multivariate analysis of COUNT score and PNI associated with in-hospital death

|  | Univariate |  | Multivariate |  |
| --- | --- | --- | --- | --- |
|  |  |  | Model 1 |  |
|  | Odds ratio<br>95% CI | P value | Odds ratio<br>95% CI | P value |
| **PNI** | 0.856<br>(0.800-0.915) | <0.001 | 0.721<br>(0.581-0.896) | 0.003 |
| Normal (high-PNI) | 1 |  | 1 |  |
| Mild (middle-PNI) | 4.727<br>(1.763-12.677) | 0.020 | 4.632<br>(0.578-37.121) | 0.149 |
| Severe (low-PNI) | 8.211<br>(3.283-20.537) | <0.001 | 24.225<br>(2.147-273.327) | 0.010 |
|  | Univariate |  | Model 2 |  |
| **CONUT score** | 1.552<br>(1.260-1.912) | <0.001 | 3.371<br>(1.124-10.106) | 0.030 |
| Normal | 1 |  | -- | -- |
| Mild | 1.545<br>(0.174-13.714) | 0.696 | -- | -- |
| Moderate | 4.435<br>(0.535-36.744) | 0.167 | -- | -- |
| Severe | 102<br>(5.480-1898.580) | 0.002 | -- | -- |

**Model 1** :The multivariate analysis of PNI associated with in-hospital death adjusted for age, sex, BMI, history of hypertension, history of chronic obstructive pulmonary disease , history of coronary heart disease ,respiratory rate, alanine transaminase, creatinine, D-dimer, white blood cell count, Lymphocyte, PCT,
**Model 2** :The multivariate analysis of COUNT score associated with in-hospital death adjusted for age, sex, BMI, history of hypertension, history of chronic obstructive pulmonary disease, history of coronary heart disease ,respiratory rate, alanine transaminase, creatinine, D-dimer, white blood cell count, Lymphocyte, PCT,

**Fig. 2.**
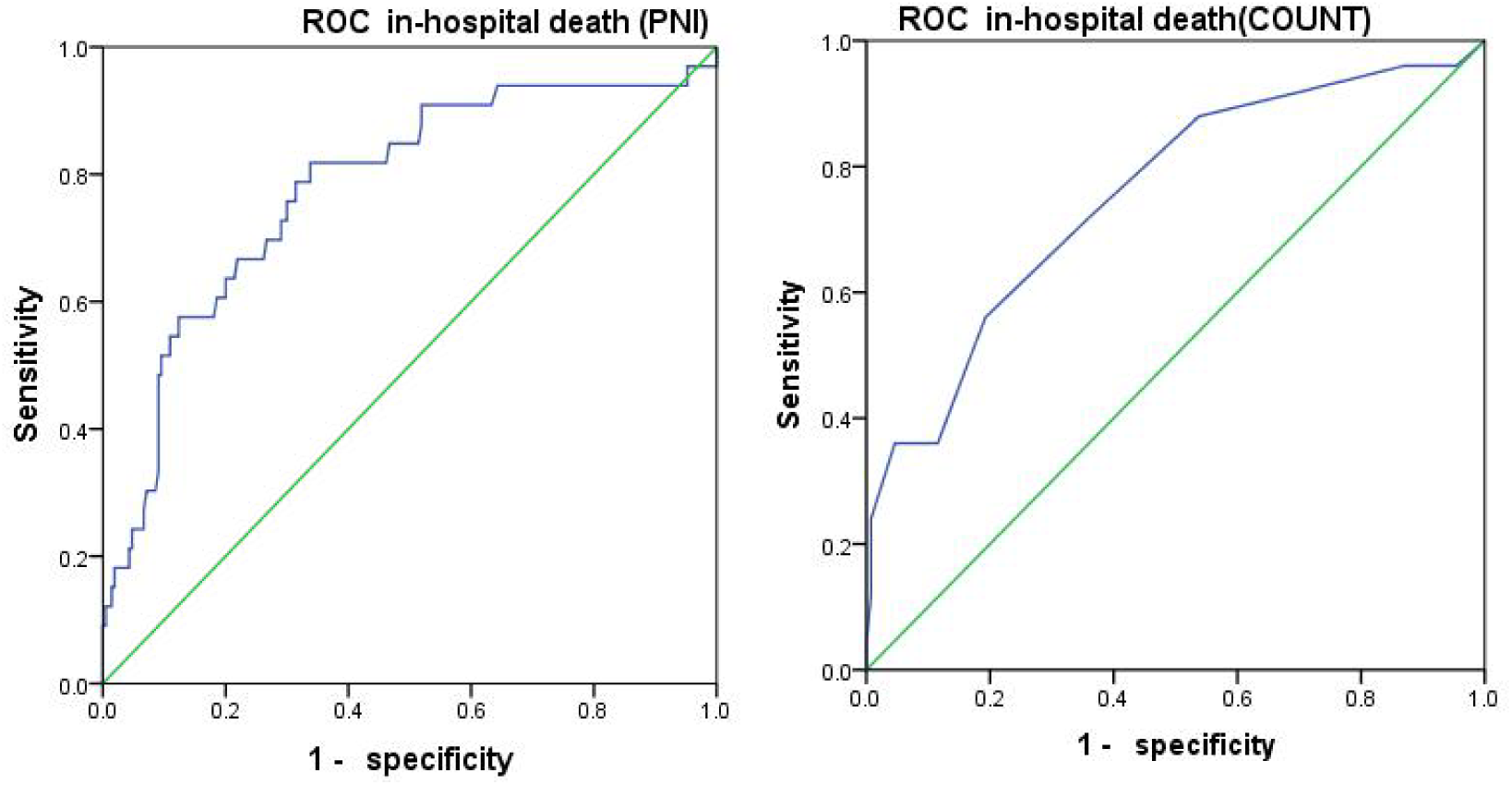
**A** PNI: sensitivity: 81.80%, specificity: 66.20%; AUC:0.778; 95%CI(0.686–0.809);p<0.001; cutoff value: 40.58; **B** TheCONUT score: sensitivity:56.00%, specificity:80.81%;AUC:0.753; 95%CI(0.644 –0.862), p<0.001; cutoff value: 5.5;

## 4. Discussion

In this study, we reported on the clinical characteristics of COVID-19 patients and the risk factors associated with death. To the author’s knowledge, this study was the first to assess the importance of two different nutritional indicators in COVID-19 patients.The COUNT score and PNI had specific statistical differences between non-survivors and survivors. However, in the multivariable logistic regression analysis, the CONUT score and PNI were the independent predictors of all-cause death, individually. According to the area under the curve of ROC,PNI was a better predictor of all-cause death than the COUNT score. In addition, according to the categorical groups of PNI, severe group was an independent predictor ofall-cause death. We could get this revelationthat the worse nutritional status, the greater risk for death. Our research found that the CONUT score and PNI might be related to some clinical features and laboratory variables.These findings pointed out that rapid assessment tools could judge the early nutritional status of COVID-19 patients and emphasized that early intervention for malnourished people could affect the prognosis.

Nutrition plays an important role in human survival and medical development.Malnutrition is caused by low nutrient feeding or malabsorption, which induced by hepatic insufficiency, anorexia, cytokine-induced high catabolism, intestinal edema. It is a common complication of many diseases[19].Malnourished patients may have the following symptoms: fatigue, weakness, low daily activities, weight loss, cognitive impairment, decreased muscle volume[20]. In this condition, the disease gradually worsens[21]. It is well known that malnutrition impairs immune function, especially cell-mediated immunity[22]. Decreased nutritional status of the body leads to decreased immunity, which directly causes or induces the occurrence of nosocomial infections, and increases the morbidity and mortality associated with infections[23].

The nutritional status assessed by the CONUT score and PNI as the prognostic indicators for patients with cardiovascular disease, malignant tumors, and acute ischemic stroke have caused the attention by researchers. And their clinical value is more comprehensive, economical, simple and practical than other tools.The CONUT score and PNI were first used to estimate the correlation between malnutrition risks and postoperative complications in patients with gastrointestinal tumors, and there was increasing evidence that they were also related to the prognosis of other malnourished malignancies and diseases[24,25,26,27].We know that the CONUT score is calculated based on 3 laboratory variables, serum albumin, lymphocytes, and total cholesterol (range 0–12, higher = worse).One study on COVID-19 confirmed that most patients had lymphopenia when they were admitted to the hospital, and that there was a decrease in albumin[28]. Studies have suggested that albumin and lymphocyte percentage might be predictors of disease severity[29]. The CONUT score weights are relatively equal between serum albumin, lymphocyte counts, and total cholesterol levels, while PNI is mainly determined by serum albumin levels and lymphocyte counts. In addition, considering that lymphocyte counts have been associated with increased disease severity. Furthermore, the lymphocytes of patients who died of COVID-19 were reported to be significantly lower than those of survivors[30].These supported our research results, which suggested that among the nutrition indicators, COUNT score and PNI wererelated to death. It has been found that some parameters are related to the prognosis of COVID-19 patients.Previously, according to the research, elderly age was an important independent predictor of death in patients with SARS and MERS[31]. Currently, COVID-19 study also confirmed that patient’s age was concerning with death. Increasing age was a predictor, which was consistent with age-related mortality study with an OR of 1.10[5]. Older age might be associated with more complications, which increased the mortality rate. In one study, predictors related to 30-day mortality included older, non-healthcare workers, pre-existing conditions, disease severity, and hospital-acquired infections[31].Interestingly, BMI was originally considered only as a way to measure body composition, but now obesity is considered a chronic inflammatory state that may affect the immune system through fat-derived inflammation[32]. COVID-19 was currently considered to have potential complex immunological damage[30]. However, in this study, BMI itself significantly differed between the death and survival groups in univariate logistics analysis model. In fact, whether BMI can be used as an evaluation index to evaluate the nutritional status of COVID-19 patients requires further study.It is unclear whether individual parameters (such as serum albumin, blood lipids, vitamins) can be used to assess the nutritional status of COVID-19 patients to predict adverse outcomes.But these single parameter indicators may also be needed to support our conclusions, especially blood lipids and albumin. The advantage of this study was that, to the author’s knowledge, this was the first study to assess the clinical significance of different nutritional indicators in patients with COVID-19. Our research will help clinicians to pay attention to the assessment of nutritional status of COVID-19 patients for early intervention and early treatment of COVID-19 patients with poor nutritional status.

## 5. Limitation

However, our study has some limitations. First, the number of patients included is relatively small. Only 155 of the 245 patients who were screened had their albumin levels, lymphocyte counts, and total cholesterol levels collected within 30 min of admission, which resulting in a relatively low sample size for COUNT scores.Second, this study was conducted at a single-center research with limited sample size. This study might have included disproportionately more patients with poor outcomes. There might also be a selection bias when identifying factors that influence the clinical outcomes.

## 6. Conclusion

In conclusion, the study has pointed out that the CONUT score and PNI were the independent risk factors for COVID-19 death. The assessment of the nutritional status of COVID-19 patients is crucial to the prognosis.

## Data Availability

YES

